# Post-exertion oxygen saturation as a prognostic factor for adverse outcome in patients attending the emergency department with suspected COVID-19: Observational cohort study

**DOI:** 10.1101/2020.08.10.20171033

**Authors:** Steve Goodacre

## Abstract

**Background:** Measurement of post-exertion oxygen saturation has been proposed to assess illness severity in suspected COVID-19 infection. We aimed to determine the accuracy of post-exertional oxygen saturation for predicting adverse outcome in suspected COVID-19.

**Methods:** We undertook an observational cohort study across 70 emergency departments during first wave of the COVID-19 pandemic in the United Kingdom. We collected data prospectively, using a standardised assessment form, and retrospectively, using hospital records, from patients with suspected COVID-19, and reviewed hospital records at 30 days for adverse outcome (death or receiving organ support). Patients with post-exertion oxygen saturation recorded were selected for this analysis.

**Results:** We analysed data from 817 patients with post-exertion oxygen saturation recorded after excluding 54 in whom measurement appeared unfeasible. The c-statistic for post-exertion change in oxygen saturation was 0.589 (95% confidence interval 0.465 to 0.713), and the positive and negative likelihood ratios of a 3% or more desaturation were respectively 1.78 (1.25 to 2.53) and 0.67 (0.46 to 0.98). Multivariable analysis showed that post-exertion oxygen saturation was not a significant predictor of adverse outcome when baseline clinical assessment was taken into account (p=0.368). Secondary analysis excluding patients in whom post-exertion measurement appeared inappropriate resulted in a c-statistic of 0.699 (0.581 to 0.817), likelihood ratios of 1.98 (1.26 to 3.10) and 0.61 (0.35 to 1.07), and some evidence of additional prognostic value on multivariable analysis (p=0.019).

**Conclusions:** Post-exertion oxygen saturation provides modest prognostic information in the assessment of patients attending the emergency department with suspected COVID-19.

**Registration:** ISRCTN registry, ISRCTN56149622, http://www.isrctn.com/ISRCTN28342533

**Key messages:** What is already known on this subject?

- Post exertional decrease in oxygen saturation can be used to predict prognosis in chronic lung diseases
- Post exertional desaturation has been proposed as a way of predicting adverse outcome in people with suspected COVID-19

What this study adds:

- Post-exertion oxygen saturation provides modest prognostic information in the assessment of patients attending the emergency department with suspected COVID-19

## Introduction

Guidelines for assessment of suspected COVID-19 recommend measurement of peripheral oxygen saturation to determine the severity of acute respiratory infection (1-3). Clinicians have noted that patients with suspected COVID-19 and a relatively normal oxygen saturation may desaturate after exertion, but the clinical importance of this finding is uncertain.

Field walking tests are commonly used to evaluate exercise capacity and assess prognosis in chronic respiratory diseases (4). The lowest arterial oxygen saturation recorded during a 6-minute walk test is an important marker of disease severity and prognosis (5). The rapid 1-minute sit-to-stand test correlates with the 6-minute walk test and the severity of lung disease (6). Exertional tests have shown desaturation in chronic obstructive lung disease (7,8) chronic interstitial lung disease (9-11) and pneumocystis carinii pneumonia (12). A modified 6-minute walk test has been proposed for use in suspected COVID-19 infection (13) but not yet evaluated, to our knowledge. A recent review of rapid exercise tests for oxygen desaturation (14) identified a number of studies, as outlined above, but found no published studies in COVID-19. The authors suggested that a 3% drop in oxygen saturation on exercise was a cause for concern, based on evidence from other conditions.

The Pandemic Respiratory Infection Emergency System Triage (PRIEST) study is a multicentre observational cohort study designed to develop and evaluate triage methods for patients with suspected COVID-19 infection. We added evaluation of post-exertion oxygen saturation to the aims of the PRIEST study in response to reports of its use in the assessment of suspected COVID-19. Our specific objective was to determine the accuracy of post-exertional oxygen saturation as a prognostic factor for 30-day adverse outcome.

## Methods

We collected data from consecutive patients presenting with suspected COVID-19 infection to 70 hospital emergency departments from 53 recruiting sites in the United Kingdom (UK). Hospitals used either prospective data collection, through a standardised assessment form for suspected COVID-19, or retrospective data collection, through research staff extracting data from hospital records onto the standardised form.

Patients were included if the assessing clinician used the standardised assessment form or recorded that the patient had suspected COVID-19 infection. The clinical diagnostic criteria used for suspected COVID-19 during the study were (1) fever (pyrexia ≥38°C) or a history of fever and (2) influenza-like illness (two or more of cough, sore throat, rhinorrhoea, limb or joint pain, headache, vomiting or diarrhoea) or severe and/or life-threatening illness suggestive of an infectious process. We did not seek consent to collect data but information about the study was provided in the ED and patients could withdraw their data at their request. Patients with multiple presentations to hospital were only included once, using data from the first presentation identified by research staff.

The population for this analysis was patients who had post-exertion oxygen saturation recorded as part of routine care. The assessing clinician made the decision to measure post-exertion oxygen saturation and determined the approach to achieving exertion. The study did not influence clinical care, so we were unable to standardise the selection of patients or the approach to measuring post-exertion oxygen saturation. Measurement could have been undertaken deliberately, by asking the patient to exercise in a specified way, or opportunistically, by recording oxygen saturation after the patient had exerted themselves for another purpose.

Research staff reviewed hospital records to identify outcomes up to 30 days after initial presentation. We defined patients who died or required respiratory, cardiovascular, or renal support as having an adverse outcome. We defined respiratory support as any intervention to protect the patient’s airway or assist their ventilation, including mechanical ventilation, non-invasive ventilation, or continuous positive airway pressure, but not supplemental oxygen alone or nebulised bronchodilators. We defined cardiovascular support as any intervention to maintain organ perfusion, including extra-corporeal membrane oxygenation, inotropic drugs, or invasive cardiovascular monitoring, but not peripheral intravenous cannulation and/or fluid administration. We defined renal support as any intervention to assist renal function, including haemofiltration, haemodialysis, or peritoneal dialysis, but not intravenous fluid administration or urinary catheterisation.

We undertook an initial descriptive analysis of the patients with post-exertion oxygen saturation recorded. This identified a number of patients for whom post-exertion oxygen saturation measurement appeared unfeasible, based on age (less than three years), performance status bed/chair bound, baseline oxygen saturation below 85%, post exertion oxygen saturation below 50%, receiving supplemental oxygen or Glasgow Coma Score less than 14. We excluded these patients from the analysis.

We examined baseline oxygen saturation, post-exertion oxygen saturation and post-exertion change in oxygen saturation (i.e. baseline minus post-exertion oxygen saturation). Analysis focused on the latter, because this indicates the additional value achieved by measuring oxygen saturation after exertion. We estimated the accuracy of each index test in terms of the sensitivity, specificity and likelihood ratios of each test across a range of thresholds for positivity, for predicting adverse outcome up to 30 days. Confidence intervals for likelihood ratios were calculated using the methods outlined in Koopman *et al*. (15) Receiving Operator Characteristic (ROC) Curves were constructed and the c-statistic (area under the ROC curve) was calculated for each index test. We did not attempt to determine an optimal threshold for positivity, because that depends upon the relative importance of sensitivity and specificity in the decision that post-exertion oxygen saturation is intended to inform. However, we decided *a priori* to highlight the performance of a 3% desaturation, as suggested by Greenhalgh *et al*. (14) Analysis was performed on patients with post-exertion oxygen saturation recorded and available 30 day outcome data, as such missing data was not imputed.

To determine whether measurement of post-exertion oxygen saturation adds prognostic information to standard respiratory assessment, we fitted a multivariable model with age, baseline oxygen saturation, respiratory rate, heart rate, asthma, other chronic respiratory illness and post-exertional oxygen saturation as covariates.

We undertook a secondary analysis that excluded patients for whom post-exertion oxygen saturation measurement appeared less appropriate, based on age (less than 16 years), performance status of limited self-care, baseline oxygen saturation less than 94%, or heart rate, respiratory rate or systolic blood pressure scoring three points on the National Early Warning Score (NEWS2). The rationale for this analysis was that local guidelines (3) recommend admission for patients with oxygen saturation less than 94% or a score of three points or more on any NEWS2 parameter. It has also been suggested that post-exertional assessment is only undertaken in a patient able to stand safely unaided and whose resting saturation is 96% or above. (14)

We planned for the PRIEST study to recruit a sample size of 20,000. The analysis presented here is a secondary analysis, so no sample size was pre-specified.

### Patient and public involvement

The Sheffield Emergency Care Forum (SECF) is a public representative group interested in emergency care research. [15] Members of SECF advised on the development of the PRIEST study and two members joined the Study Steering Committee. Patients were not involved in the recruitment to and conduct of the study. We are unable to disseminate the findings to study participants directly.

## Results

The PRIEST study recruited 22485 patients across 70 hospitals between 26 March 2020 and 28 May 2020, of whom 39 requested withdrawal of their data. We identified 874 patients who had post-exertion oxygen saturation recorded and excluded 57 in whom measurement appeared unfeasible, leaving 817 for analysis. Adverse outcome occurred in 30 participants (3.7%), of these nine died, 22 had respiratory support, five had cardiovascular support and four renal support.

Supplemental Figure S1 shows the flow of patients through the study, and Supplemental Table S1 shows the characteristics of the whole PRIEST cohort and the characteristics of those included in this analysis. Participants in this analysis were younger, more likely to have unrestricted performance status, less likely to have any comorbidities, tended to have more normal baseline physiology and had a much lower rate of adverse outcome.

Table 1 compares the baseline oxygen saturation, post-exertion oxygen saturation and post-exertion change between those with and without an adverse outcome. Post-exertion oxygen saturation tended to be lower than baseline oxygen saturation and show a greater decrease in those who suffered adverse outcome (2.9% versus 1.9% mean decrease). However, Figure 1 show that oxygen saturations increased post-exertion in a proportion of cases and there was considerable overlap between those with and without adverse outcome. Supplemental Figures S2 and S3 show overlayed histograms for baseline and post-exertion oxygen saturation.

**Figure 1:**
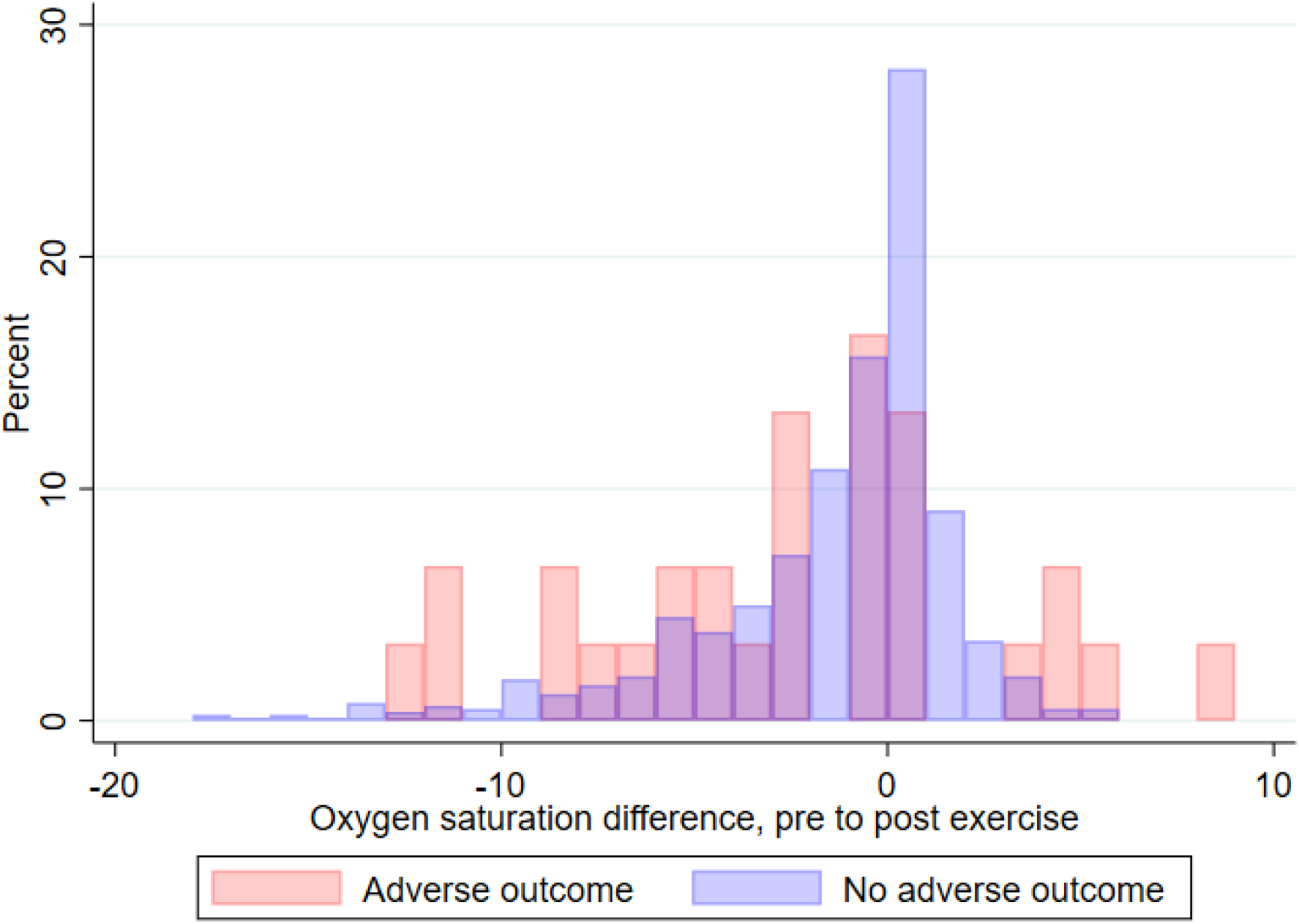
Overlayed histogram comparing post-exertion change in oxygen saturation from baseline between patients with and without adverse outcome (N=813)

**Table 1:**
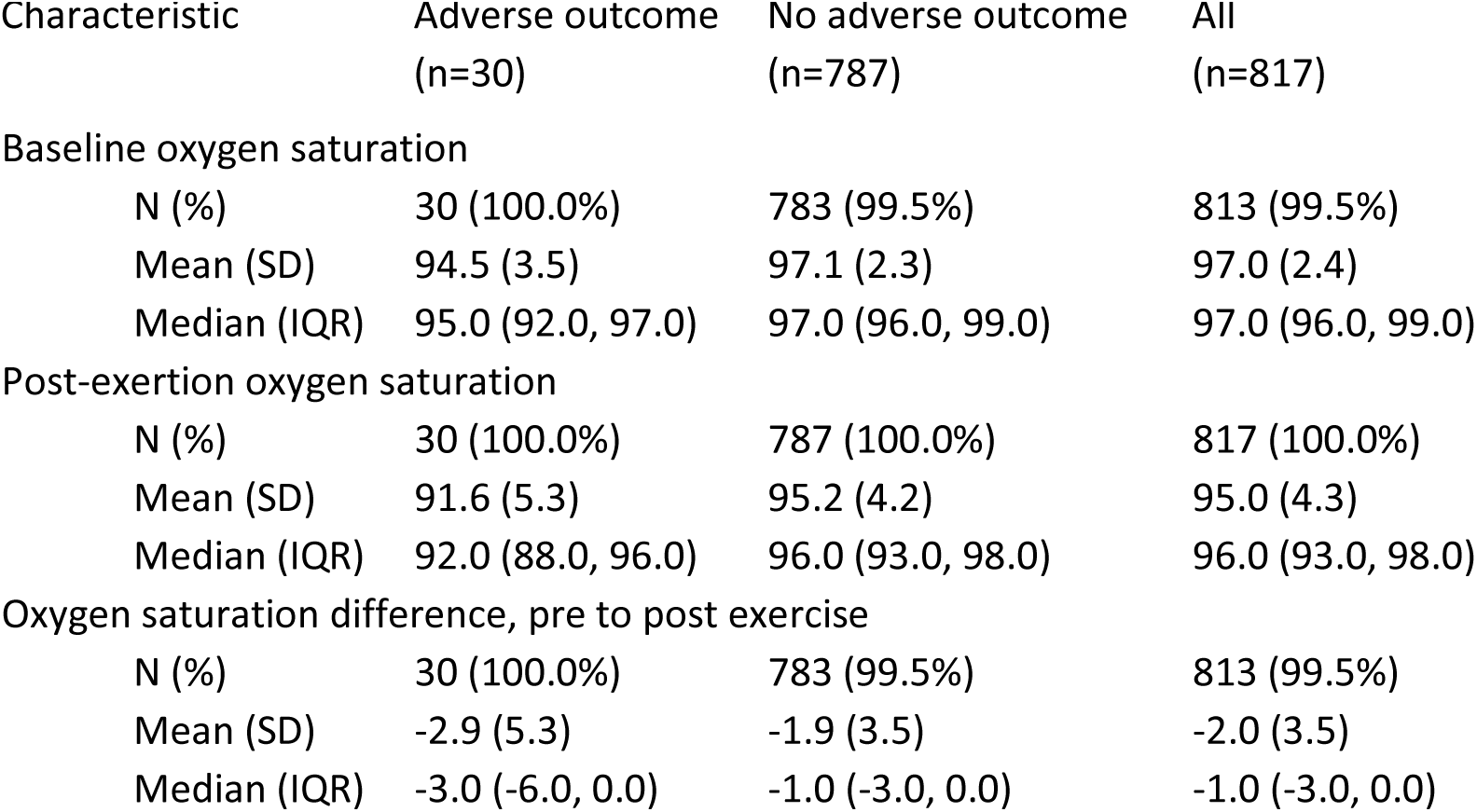
Comparison of index tests summary statistics between those with and without adverse outcome.

| Characteristic | Adverse outcome<br>(n=30) | No adverse outcome<br>(n=787) | All<br>(n=817) |
| --- | --- | --- | --- |
| Baseline oxygen saturation |  |  |  |
| N (%) | 30 (100.0%) | 783 (99.5%) | 813 (99.5%) |
| Mean (SD) | 94.5 (3.5) | 97.1 (2.3) | 97.0 (2.4) |
| Median (IQR) | 95.0 (92.0, 97.0) | 97.0 (96.0, 99.0) | 97.0 (96.0, 99.0) |
| Post-exertion oxygen saturation |  |  |  |
| N (%) | 30 (100.0%) | 787 (100.0%) | 817 (100.0%) |
| Mean (SD) | 91.6 (5.3) | 95.2 (4.2) | 95.0 (4.3) |
| Median (IQR) | 92.0 (88.0, 96.0) | 96.0 (93.0, 98.0) | 96.0 (93.0, 98.0) |
| Oxygen saturation difference, pre to post exercise |  |  |  |
| N (%) | 30 (100.0%) | 783 (99.5%) | 813 (99.5%) |
| Mean (SD) | -2.9 (5.3) | -1.9 (3.5) | -2.0 (3.5) |
| Median (IQR) | -3.0 (-6.0, 0.0) | -1.0 (-3.0, 0.0) | -1.0 (-3.0, 0.0) |

Figure 2 shows the ROC curves for baseline oxygen saturation, post-exertion oxygen saturation and post-exertion change in oxygen saturation. The c-statistic of 0.589 for post-exertion change indicates poor discriminant value, partly due to post-exertion increases in oxygen saturation showing some association with adverse outcome. Table 2 reports sensitivity, specificity and likelihood ratios for thresholds of post-exertion decrease in oxygen saturation (i.e. change less than zero). The positive and negative likelihood ratios of a post-exertional desaturation of 3% or more were 1.78 and 0.67 respectively, suggesting that this finding provides a small amount of additional information in prognostic assessment. Supplemental tables S2 and S3 show the diagnostic parameters for baseline and post-exertion oxygen saturation respectively.

**Figure 2:**
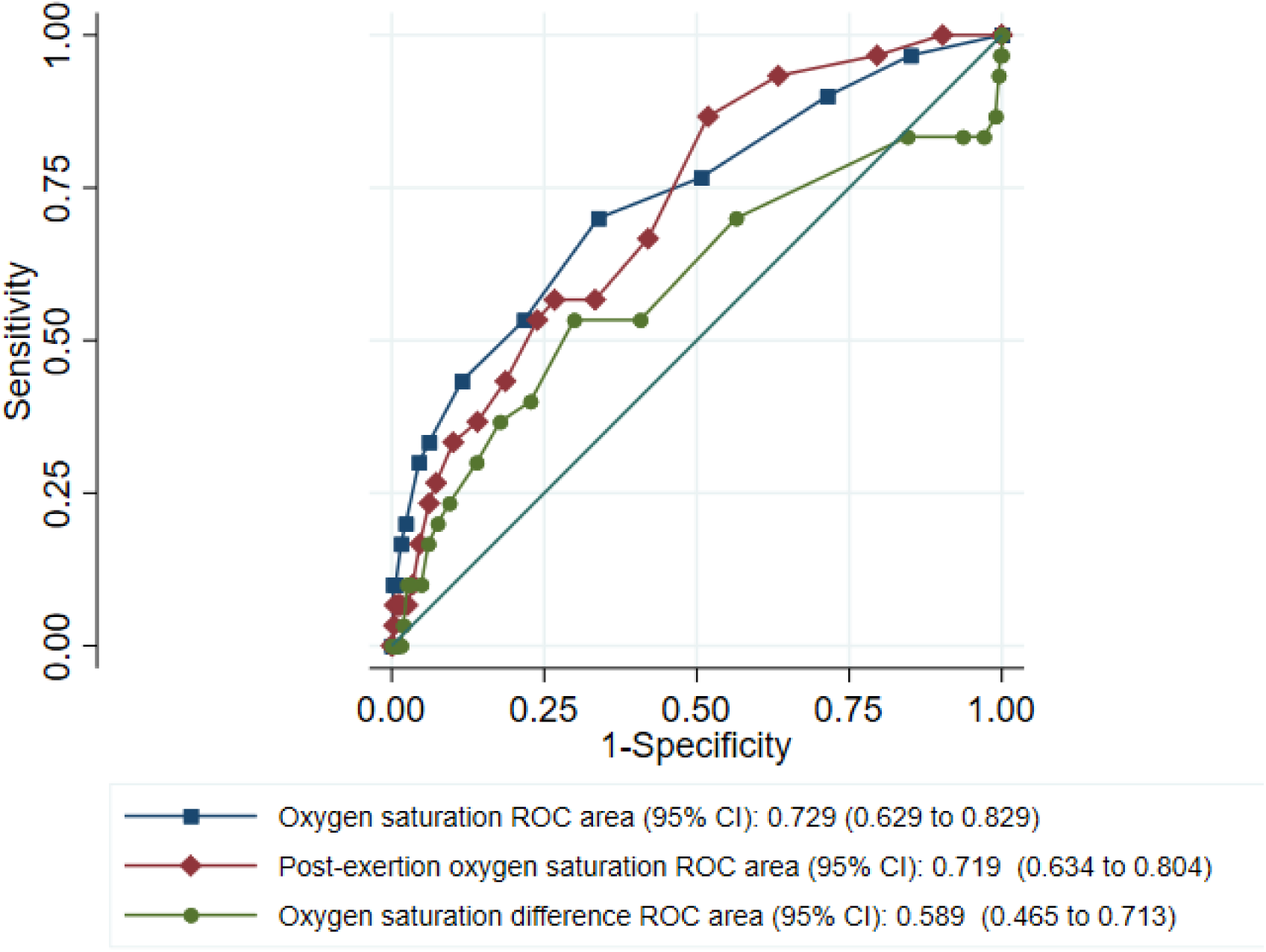
ROC curves showing index test accuracies for predicting adverse outcome, primary analysis (n=817)

**Table 2:** Accuracy of post-exertion change in oxygen saturation from baseline at a range of thresholds for positivity, primary analysis (N=813)

| Threshold | Sensitivity<br>(95% CI) | Specificity<br>(95% CI) | Positive<br>likelihood ratio<br>(95% CI) | Negative<br>likelihood ratio<br>(95% CI) |
| --- | --- | --- | --- | --- |
|  | 70.0 | 43.6 | 1.24 | 0.69 |
| <=-1 | (50.6 to 85.3) | (40.0 to 47.1) | (0.97 to 1.58) | (0.40 to 1.20) |
|  | 53.3 | 59.3 | 1.31 | 0.79 |
| <=-2 | (34.3 to 71.7) | (55.7 to 62.7) | (0.93 to 1.85) | (0.54 to 1.16) |
|  | 53.3 | 70.1 | 1.78 | 0.67 |
| <=-3 | (34.3 to 71.7) | (66.8 to 73.3) | (1.25 to 2.53) | (0.46 to 0.98) |
|  | 40.0 | 77.3 | 1.76 | 0.78 |
| <=-4 | (22.7 to 59.4) | (74.2 to 80.2) | (1.11 to 2.78) | (0.58 to 1.05) |
|  | 36.7 | 82.2 | 2.07 | 0.77 |
| <=-5 | (19.9 to 56.1) | (79.4 to 84.9) | (1.26 to 3.39) | (0.59 to 1.01) |
|  | 30.0 | 86.1 | 2.16 | 0.81 |
| <=-6 | (14.7 to 49.4) | (83.5 to 88.4) | (1.22 to 3.83) | (0.64 to 1.03) |
|  | 23.3 | 90.5 | 2.47 | 0.85 |
| <=-7 | (9.9 to 42.3) | (88.3 to 92.5) | (1.25 to 4.89) | (0.70 to 1.04) |
|  | 20.0 | 92.5 | 2.65 | 0.87 |
| <=-8 | (7.7 to 38.6) | (90.4 to 94.2) | (1.24 to 5.65) | (0.73 to 1.04) |
|  | 16.7 | 94.0 | 2.78 | 0.89 |
| <=-9 | (5.6 to 34.7) | (92.1 to 95.6) | (1.19 to 6.48) | (0.76 to 1.05) |
|  | 10.0 | 95.1 | 2.06 | 0.95 |
| <=-10 | (2.1 to 26.5) | (93.4 to 96.5) | (0.67 to 6.30) | (0.84 to 1.07) |

Figure 3 and Table 3 show comparable results for the secondary analysis excluding cases where post-exertion oxygen saturation measurement appeared less appropriate. The c-statistic of 0.699 for post-exertion change in oxygen saturation indicates better discriminant value in this group. This may be explained by exclusion of patients with lower baseline oxygen saturations who had more potential to show a post-exertion change. The positive and negative likelihood ratios of a post-exertional decrease in oxygen saturation of 3% or more were 1.98 and 0.61 respectively. Supplemental tables S4 and S5 show the diagnostic parameters for baseline and post-exertion oxygen saturation respectively for the secondary analysis.

**Fig 3:**
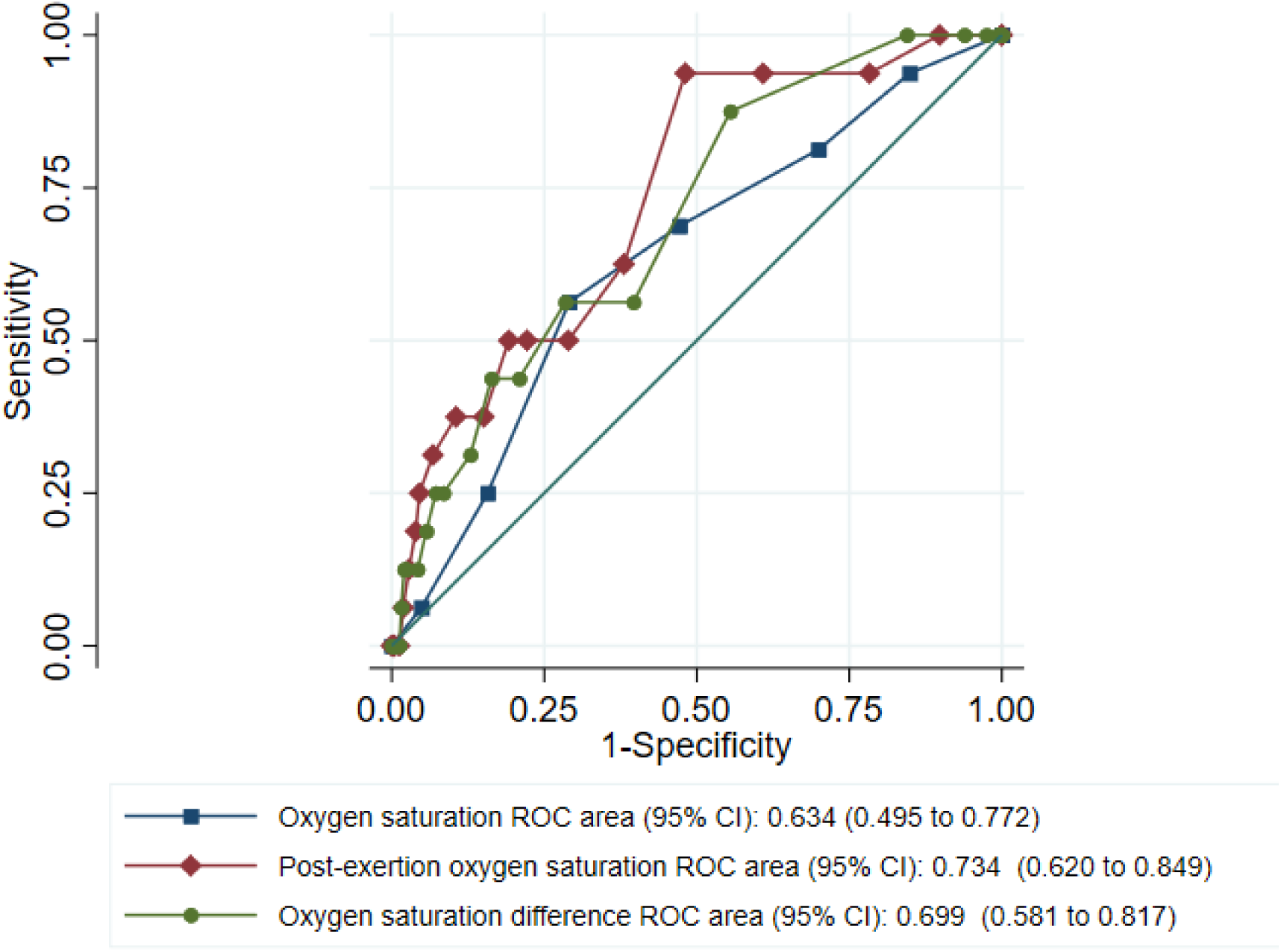
ROC curves showing index test accuracies for predicting adverse outcome, secondary analysis (N=655)

**Table 3:** Accuracy of post-exertion change in oxygen saturation from baseline at a range of thresholds for positivity, secondary analysis (N=652)

| Threshold | Sensitivity<br>(95% CI) | Specificity<br>(95% CI) | Positive<br>likelihood ratio<br>(95% CI) | Negative<br>likelihood ratio<br>(95% CI) |
| --- | --- | --- | --- | --- |
|  | 87.5 | 44.5 | 1.58 | 0.28 |
| <=-1 | (61.7 to 98.4) | (40.6 to 48.5) | (1.30 to 1.93) | (0.08 to 1.03) |
|  | 56.3 | 60.4 | 1.42 | 0.72 |
| <=-2 | (29.9 to 80.2) | (56.5 to 64.2) | (0.91 to 2.21) | (0.41 to 1.26) |
|  | 56.3 | 71.5 | 1.98 | 0.61 |
| <=-3 | (29.9 to 80.2) | (67.9 to 75.0) | (1.26 to 3.10) | (0.35 to 1.07) |
|  | 43.8 | 79.1 | 2.09 | 0.71 |
| <=-4 | (19.8 to 70.1) | (75.7 to 82.2) | (1.18 to 3.72) | (0.46 to 1.10) |
|  | 43.8 | 83.6 | 2.68 | 0.67 |
| <=-5 | (19.8 to 70.1) | (80.5 to 86.4) | (1.50 to 4.80) | (0.43 to 1.03) |
|  | 31.3 | 87.1 | 2.42 | 0.79 |
| <=-6 | (11.0 to 58.7) | (84.3 to 89.6) | (1.14 to 5.15) | (0.57 to 1.10) |
|  | 25.0 | 91.5 | 2.94 | 0.82 |
| <=-7 | (7.3 to 52.4) | (89.1 to 93.6) | (1.21 to 7.13) | (0.62 to 1.09) |
|  | 25.0 | 92.8 | 3.46 | 0.81 |
| <=-8 | (7.3 to 52.4) | (90.5 to 94.7) | (1.42 to 8.45) | (0.61 to 1.08) |
|  | 18.8 | 94.3 | 3.31 | 0.86 |
| <=-9 | (4.0 to 45.6) | (92.2 to 96.0) | (1.14 to 9.63) | (0.68 to 1.09) |
|  | 12.5 | 95.8 | 2.94 | 0.91 |
| <=-10 | (1.6 to 38.3) | (93.9 to 97.2) | (0.76 to 11.32) | (0.76 to 1.10) |

In the multivariable model on the primary analysis cohort, post-exertional oxygen saturation did not add prognostic value over other factors in the model (p-value for model coefficient 0.368, likelihood ratio test for model with and without post-exertion oxygen saturation 0.78, p=0.376). For the secondary analysis, post-exertional oxygen saturation added prognostic value over other factors (p-value for model coefficient 0.019, likelihood ratio test for model with and without post-exertion oxygen saturation 4.82, p=0.078).

## Discussion

Our findings suggest that measurement of post-exertion oxygen saturation adds little to clinical assessment of suspected COVID-19 in the emergency department. The likelihood ratios suggest that a desaturation of 3% or more provides a small amount of prognostic information, the c-statistic of 0.589 for post-exertion change suggests little discriminant value, and multivariate analysis suggest that post-exertion oxygen saturation measurement does not add prognostic value once baseline measurements are taken into account. Secondary analysis suggested better discriminant value and some additional prognostic value when testing was limited to more appropriate cases, but likelihood ratios still suggested that a desaturation of 3% or more still provided only modest prognostic information.

The observation that oxygen saturation increased post-exertion, and that some people with adverse outcome showed an increase, may seem surprising, but is probably explained by random variation. Oxygen saturation varies randomly from one measurement to the next and this variation is likely to be greater in sicker patients with baseline hypoxia. Thus we might expect greater variation in oxygen saturation to show some association with adverse outcome.

Our findings suggest that measurement of post-exertion oxygen saturation provides little additional prognostic information in selected patients with suspected COVID-19. Greenhalgh *et al* suggested using a desaturation of at least 3% to identify cause for concern in selected patients who are well enough for out of hospital management. Our findings suggest that a 3% desaturation indicates small increase in the likelihood of adverse outcome. Further research could determine whether a more systematic and rigorously controlled approach to post-exertion oxygen saturation measurement can result in more useful prognostic information. The feasibility of such research may be limited by low event rates in people who are able to undertake formal post-exertion measurement of oxygen saturation.

Our study consisted of a clinically relevant population and was recruited across a wide range of settings, but evaluation of post-exertion oxygen saturation was a post hoc secondary analysis and the study was not designed specifically for this purpose. We are unable to say how patients were selected for measurement of post-exertion oxygen saturation, and the method for undertaking exertion was not standardised or recorded. We excluded 57 patients from analysis for whom post-exertion oxygen saturation measurement appeared unfeasible, and excluded a further 162 from secondary analysis for whom measurement appeared less appropriate. These cases may reflect opportunistic oxygen saturation measurement after exertion, such as on attempting to mobilise, but we cannot exclude the possibility of data recording errors. Only 874 out of 22446 patients had post-exertion oxygen saturation recorded. This may reflect limited awareness and use of post-exertion oxygen saturation, but may also reflect severity of illness in the emergency department population. Measurement of post-exertion oxygen saturation is only likely to be feasible and clinically indicated in those with milder illness. The relatively small number of adverse outcomes (N=30) limited the precision of our estimates of sensitivity and power to undertake multivariable analysis.

In summary, measuring post-exertion oxygen saturation provides little prognostic information in the assessment of patients attending the emergency department with suspected COVID-19.

## Data Availability

Anonymised data are available from the corresponding author upon reasonable request (contact details on first page). The Confidentiality Advisory Group of the Health Research Authority will need to consider any requests for data to be used for purposes other than those specified in our application, so a data request should be accompanied by explanation of the purpose of the request and justification of the public benefit. We also recommend including a pre-specified plan of analysis.

## Ethical approval

The North West - Haydock Research Ethics Committee gave a favourable opinion on the PAINTED study on 25 June 2012 (reference 12/NW/0303) and on the updated PRIEST study on 23rd March 2020. The Confidentiality Advisory Group of the Health Research Authority granted approval to collect data without patient consent in line with Section 251 of the National Health Service Act 2006.

## Acknowledgements

We thank Katie Ridsdale for clerical assistance with the study, Erica Wallis (Sponsor representative, all members of the PRIEST Research Team (Appendix 1), all members of the Study Steering Committee (Appendix 2) and the site research teams who delivered the data for the study (Appendix 3), and the research team at the University of Sheffield past and present (Appendix 4).

## Author contributions

SG, AB, KC, CF, TH, FL, ALe, IM and DW conceived and designed the study. BT, KB, ALo, SW, RS, JS, SC, ES, JH and EY acquired the data. EL, LS, SG, BT, KB and CM analysed the data. SG, AB, KC, CF, TH, FL, ALe, IM, DW, EL, LS, SG, BT, KB and CM interpreted the data. All authors contributed to drafting the manuscript. SG takes responsibility for the paper as a whole.

---

**Supplemental Table S1:** Characteristics of the PRIEST cohort and the cohort included in this analysis.

| Characteristic | All<br>(n=22446) | With post-exertion oxygen<br>saturation<br>(n=817) |
| --- | --- | --- |
| Sex |  |  |
| Male | 11035 (49.2%) | 369 (45.2%) |
| Female | 11200 (49.9%) | 442 (54.1%) |
| Missing | 211 (0.9%) | 6 (0.7%) |
| Ethnicity |  |  |
| UK\Irish\other white | 15198 (67.7%) | 475 (58.1%) |
| Asian | 1150 (5.1%) | 67 (8.2%) |
| Black/African/Caribbean | 692 (3.1%) | 47 (5.8%) |
| Mixed/multiple ethnic groups | 328 (1.5%) | 18 (2.2%) |
| Other | 570 (2.5%) | 47 (5.8%) |
| Missing | 4508 (20.1%) | 163 (20.0%) |
| Performance status |  |  |
| Unrestricted normal activity | 11917 (53.1%) | 643 (78.7%) |
| Limited strenuous activity, can do light | 2393 (10.7%) | 83 (10.2%) |
| Limited activity, can self care | 2790 (12.4%) | 36 (4.4%) |
| Limited self care | 2662 (11.9%) | 11 (1.3%) |
| Bed/chair bound, no self care | 1510 (6.7%) | 0 (0.0%) |
| Missing | 1174 (5.2%) | 44 (5.4%) |
| Age |  |  |
| N (%) | 22439 (100.0%) | 817 (100.0%) |
| Mean (SD) | 58.4 (24.2) | 48.4 (16.1) |
| Median (IQR) | 62.0 (43.0, 78.0) | 47.0 (36.0, 59.0) |
| Respiratory rate |  |  |
| N (%) | 21844 (97.3%) | 796 (97.4%) |
| Mean (SD) | 23.9 (7.7) | 20.8 (5.3) |
| Median (IQR) | 22.0 (18.0, 28.0) | 20.0 (18.0, 23.0) |
| Pulse Rate |  |  |
| N (%) | 21967 (97.9%) | 809 (99.0%) |
| Mean (SD) | 97.8 (24.5) | 90.6 (18.1) |
| Median (IQR) | 95.0 (81.0, 111.0) | 90.0 (78.0, 102.0) |
| Temperature |  |  |
| N (%) | 21741 (96.9%) | 799 (97.8%) |
| Mean (SD) | 37.2 (1.1) | 37.0 (0.8) |
| Median (IQR) | 37.0 (36.5, 37.9) | 36.8 (36.5, 37.3) |
| Systolic BP |  |  |
| N (%) | 20698 (92.2%) | 793 (97.1%) |
| Mean (SD) | 134.2 (25.0) | 136.5 (21.3) |
| Median (IQR) | 132.0 (117.0, 149.0) | 134.0 (122.0, 148.0) |
| Diastolic BP |  |  |
| N (%) | 20601 (91.8%) | 788 (96.5%) |
| Mean (SD) | 78.0 (16.2) | 82.6 (12.9) |
| Median (IQR) | 78.0 (68.0, 88.0) | 82.0 (74.5, 90.0) |
| Oxygen saturation |  |  |
| N (%) | 22155 (98.7%) | 813 (99.5%) |
| Mean (SD) | 94.9 (6.6) | 97.0 (2.4) |
| Median (IQR) | 96.0 (94.0, 98.0) | 97.0 (96.0, 99.0) |
| Medical History |  |  |
| No Chronic disease | 7077 (31.5%) | 406 (49.7%) |
| Heart Disease | 4723 (21.0%) | 66 (8.1%) |
| Renal impairment | 1944 (8.7%) | 27 (3.3%) |
| Steroid therapy | 564 (2.5%) | 13 (1.6%) |
| Asthma | 3492 (15.6%) | 135 (16.5%) |
| Diabetes | 4132 (18.4%) | 73 (8.9%) |
| Active malignancy | 1124 (5.0%) | 14 (1.7%) |
| Immunosuppression | 646 (2.9%) | 30 (3.7%) |
| Other chronic lung disease | 3795 (16.9%) | 81 (9.9%) |
| Hypertension | 6440 (28.7%) | 155 (19.0%) |
| Medical history missing | 1104 (4.9%) | 50 (6.1%) |
| Adverse outcome |  |  |
| Any | 4698 (20.9%) | 30 (3.7%) |
| Death | 3324 (14.8%) | 9 (1.1%) |
| Respiratory support | 1962 (8.7%) | 22 (2.7%) |
| Cardiovascular support | 525 (2.3%) | 5 (0.6%) |
| Renal support | 220 (1.0%) | 4 (0.5%) |

**Supplemental Table S2:** Accuracy of baseline oxygen saturation at a range of thresholds for positivity, primary analysis (N=813)

| Threshold | Sensitivity<br>(95% CI) | Specificity<br>(95% CI) | Positive likelihood<br>ratio (95% CI) | Negative likelihood<br>ratio (95% CI) |
| --- | --- | --- | --- | --- |
| <=99 | 96.7 (82.8 to 99.9) | 14.9 (12.5 to 17.6) | 1.14 (1.06 to 1.23) | 0.22 (0.03 to 1.52) |
| <=98 | 90.0 (73.5 to 97.9) | 28.6 (25.5 to 31.9) | 1.26 (1.11 to 1.43) | 0.35 (0.12 to 1.03) |
| <=97 | 76.7 (57.7 to 90.1) | 49.3 (45.7 to 52.9) | 1.51 (1.23 to 1.86) | 0.47 (0.24 to 0.90) |
| <=96 | 70.0 (50.6 to 85.3) | 66.2 (62.7 to 69.5) | 2.07 (1.61 to 2.67) | 0.45 (0.26 to 0.78) |
| <=95 | 53.3 (34.3 to 71.7) | 78.3 (75.2 to 81.1) | 2.46 (1.72 to 3.53) | 0.60 (0.41 to 0.88) |
| <=94 | 43.3 (25.5 to 62.6) | 88.5 (86.1 to 90.7) | 3.77 (2.40 to 5.93) | 0.64 (0.47 to 0.88) |
| <=93 | 33.3 (17.3 to 52.8) | 93.9 (92.0 to 95.4) | 5.44 (3.06 to 9.67) | 0.71 (0.55 to 0.92) |
| <=92 | 30.0 (14.7 to 49.4) | 95.5 (93.8 to 96.9) | 6.71 (3.55 to 12.67) | 0.73 (0.58 to 0.92) |
| <=91 | 20.0 (7.7 to 38.6) | 97.7 (96.4 to 98.6) | 8.70 (3.72 to 20.33) | 0.82 (0.69 to 0.98) |
| <=90 | 16.7 (5.6 to 34.7) | 98.5 (97.3 to 99.2) | 10.87 (4.09 to 28.89) | 0.85 (0.72 to 1.00) |

**Supplemental Table S3:** Accuracy of post-exertion oxygen saturation at a range of thresholds for positivity, primary analysis (N=817)

| Threshold | Sensitivity<br>(95% CI) | Specificity<br>(95% CI) | Positive likelihood<br>ratio (95% CI) | Negative likelihood<br>ratio (95% CI) |
| --- | --- | --- | --- | --- |
| <=99 | 100 (88.4 to 100) | 9.8 (7.8 to 12.1) | 1.11 | 0.00 |
| <=98 | 96.7 (82.8 to 99.9) | 20.5 (17.7 to 23.4) | 1.22 (1.13 to 1.32) | 0.16 (0.02 to 1.10) |
| <=97 | 93.3 (77.9 to 99.2) | 36.7 (33.3 to 40.2) | 1.47 (1.32 to 1.64) | 0.18 (0.05 to 0.69) |
| <=96 | 86.7 (69.3 to 96.2) | 48.4 (44.9 to 52.0) | 1.68 (1.44 to 1.96) | 0.28 (0.11 to 0.70) |
| <=95 | 66.7 (47.2 to 82.7) | 58.2 (54.7 to 61.7) | 1.59 (1.22 to 2.07) | 0.57 (0.34 to 0.95) |
| <=94 | 56.7 (37.4 to 74.5) | 66.8 (63.4 to 70.1) | 1.71 (1.23 to 2.37) | 0.65 (0.43 to 0.98) |
| <=93 | 56.7 (37.4 to 74.5) | 73.4 (70.2 to 76.5) | 2.13 (1.53 to 2.97) | 0.59 (0.39 to 0.89) |
| <=92 | 53.3 (34.3 to 71.7) | 76.2 (73.1 to 79.2) | 2.24 (1.57 to 3.20) | 0.61 (0.42 to 0.90) |
| <=91 | 43.3 (25.5 to 62.6) | 81.4 (78.6 to 84.1) | 2.34 (1.52 to 3.61) | 0.70 (0.51 to 0.96) |
| <=90 | 36.7 (19.9 to 56.1) | 86.0 (83.4 to 88.4) | 2.62 (1.59 to 4.32) | 0.74 (0.56 to 0.97) |

**Table S4:**
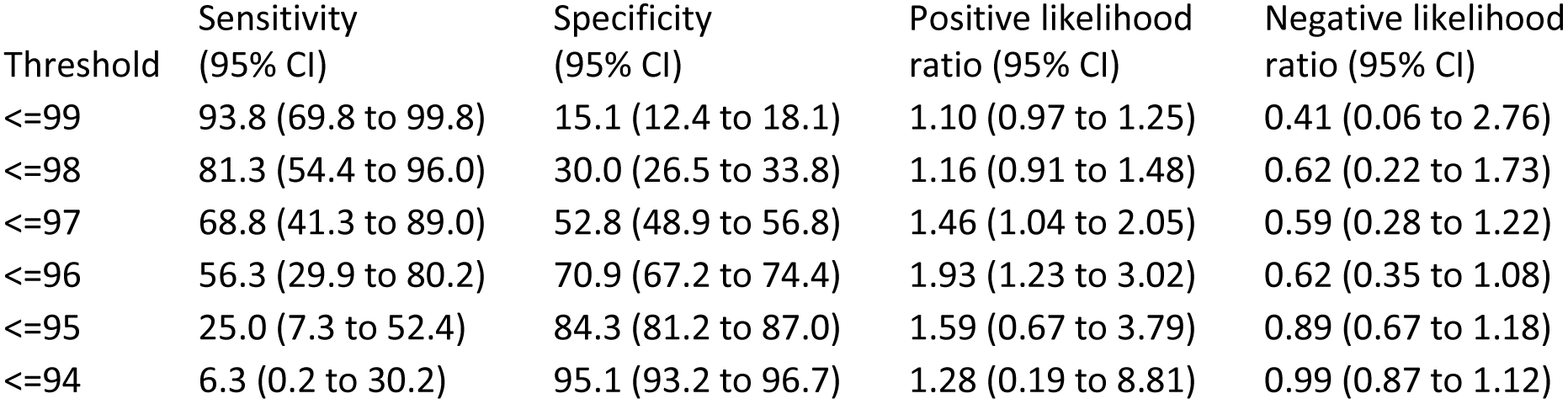
Accuracy of baseline oxygen saturation at a range of thresholds for positivity, secondary analysis (N=652)

| Threshold | Sensitivity<br>(95% CI) | Specificity<br>(95% CI) | Positive likelihood<br>ratio (95% CI) | Negative likelihood<br>ratio (95% CI) |
| --- | --- | --- | --- | --- |
| <=99 | 93.8 (69.8 to 99.8) | 15.1 (12.4 to 18.1) | 1.10 (0.97 to 1.25) | 0.41 (0.06 to 2.76) |
| <=98 | 81.3 (54.4 to 96.0) | 30.0 (26.5 to 33.8) | 1.16 (0.91 to 1.48) | 0.62 (0.22 to 1.73) |
| <=97 | 68.8 (41.3 to 89.0) | 52.8 (48.9 to 56.8) | 1.46 (1.04 to 2.05) | 0.59 (0.28 to 1.22) |
| <=96 | 56.3 (29.9 to 80.2) | 70.9 (67.2 to 74.4) | 1.93 (1.23 to 3.02) | 0.62 (0.35 to 1.08) |
| <=95 | 25.0 (7.3 to 52.4) | 84.3 (81.2 to 87.0) | 1.59 (0.67 to 3.79) | 0.89 (0.67 to 1.18) |
| <=94 | 6.3 (0.2 to 30.2) | 95.1 (93.2 to 96.7) | 1.28 (0.19 to 8.81) | 0.99 (0.87 to 1.12) |

**Table S5:**
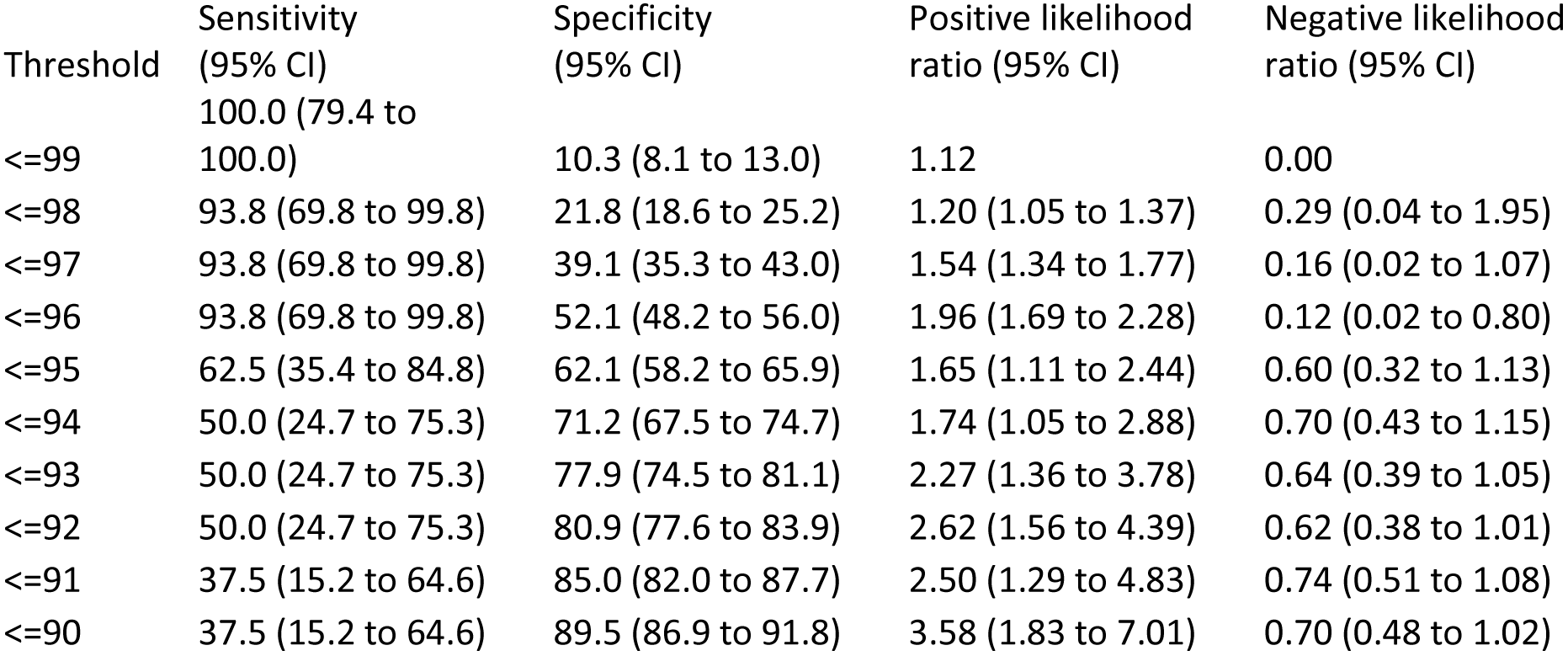
Accuracy of post-exertion oxygen saturation at a range of thresholds for positivity, secondary analysis.

**Figure S1:**
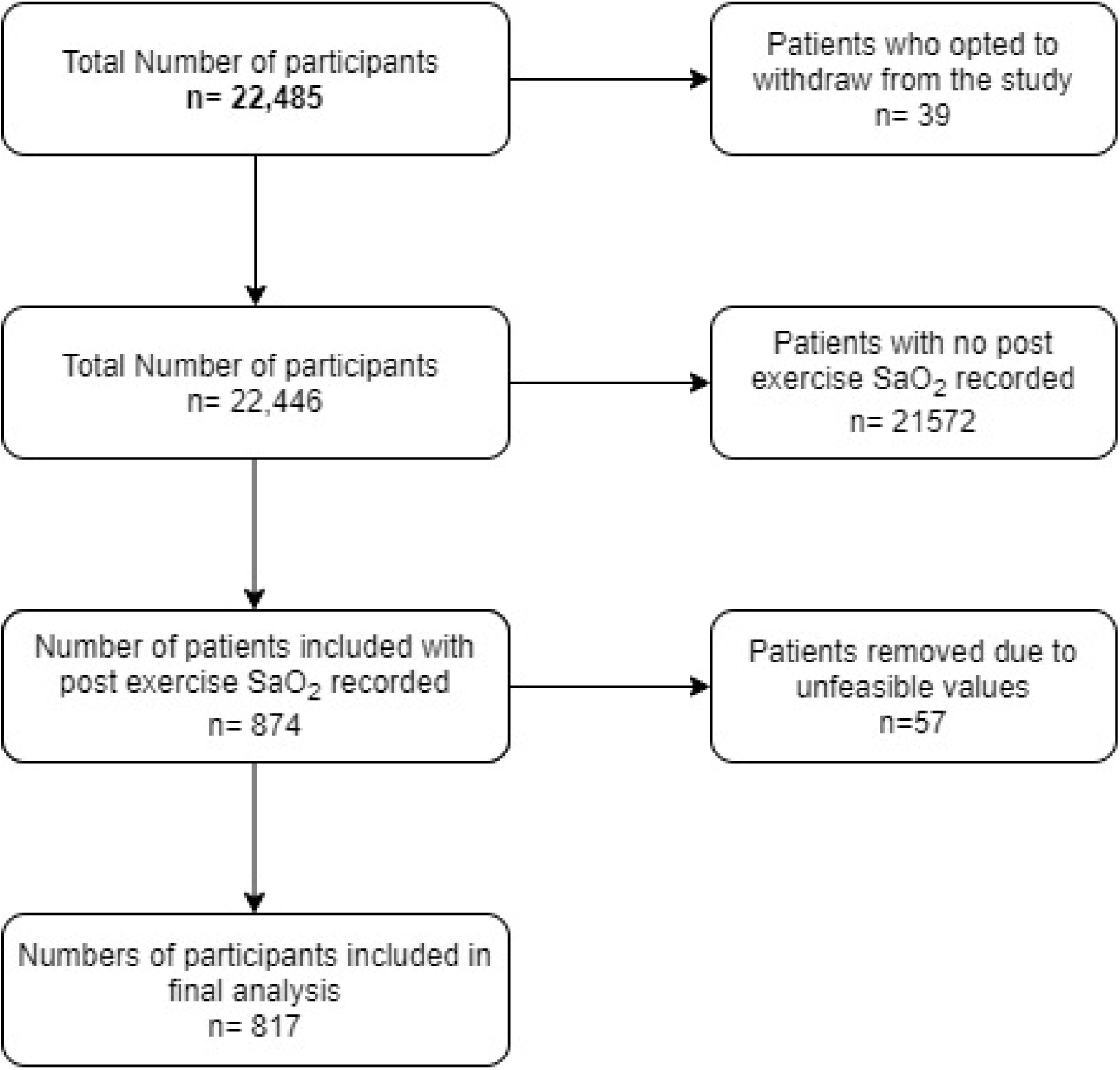
Flow of patients through the study.

**Figure S2:**
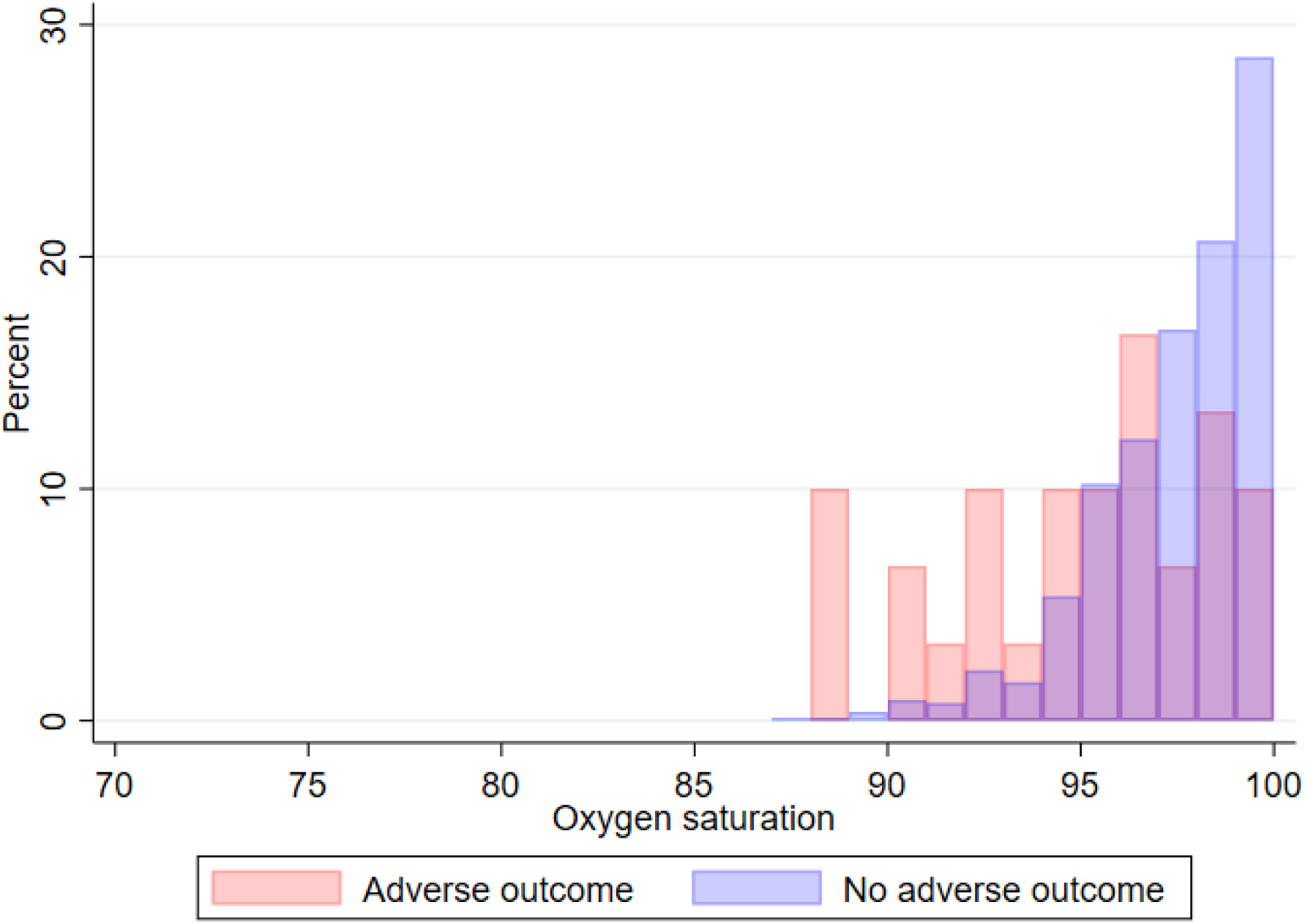
Overlayed histogram comparing baseline oxygen saturation between patients with and without adverse outcome (N=813)

**Figure S3:**
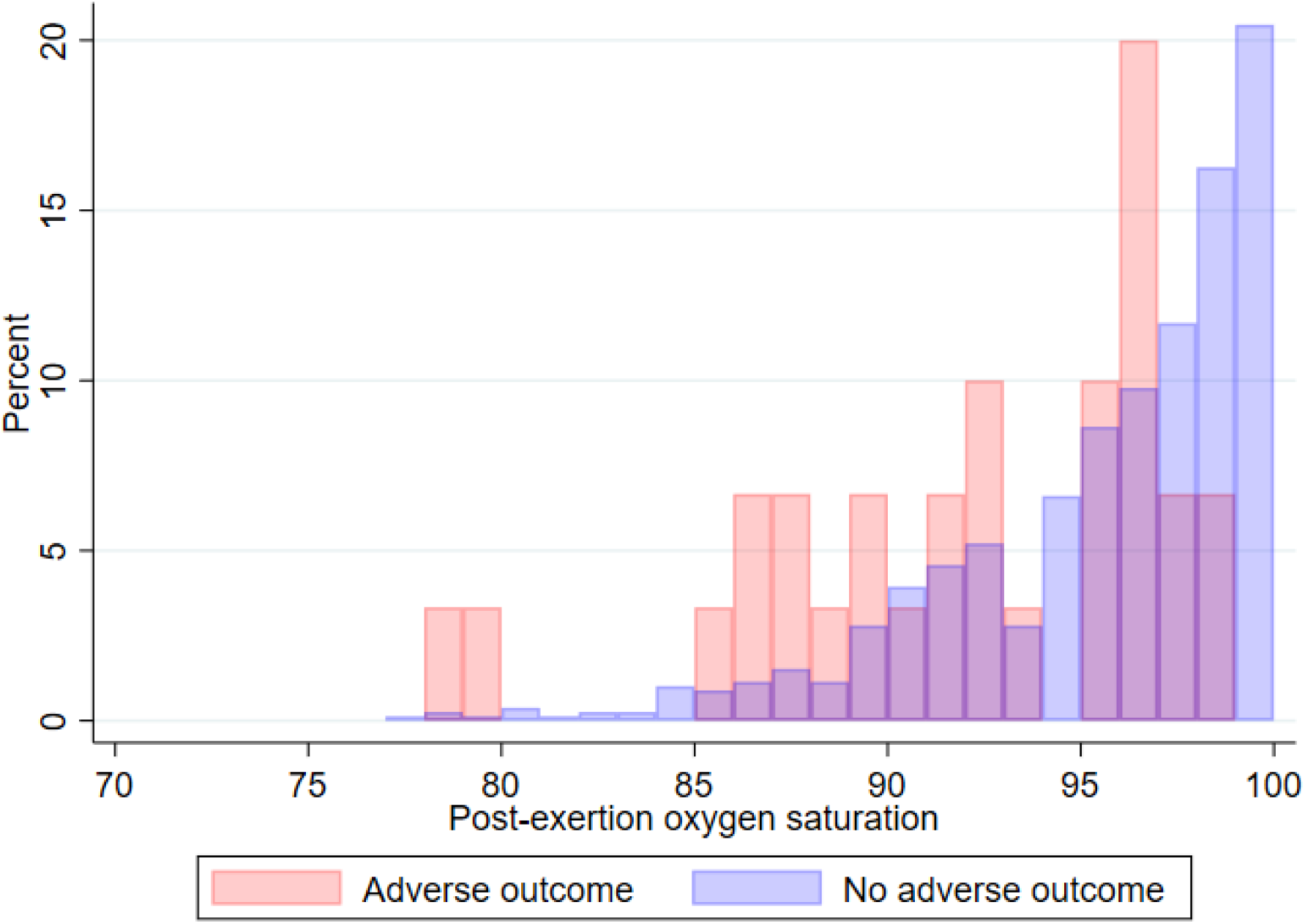
Overlayed histogram comparing post-exertion oxygen saturation between patients with and without adverse outcome (N=817)

